# Developing visit usage recommendations for outpatient rehabilitation after total knee arthroplasty: A Delphi study

**DOI:** 10.1101/2022.11.29.22282593

**Authors:** Jeremy Graber, Laura Churchill, Tamara Struessel, Shane O’Malley, Michael Bade, Jennifer Stevens-Lapsley

## Abstract

**Objective:** No consensus recommendations exist for rehabilitation visit usage (i.e., visit timing, visit frequency, duration of care) after total knee arthroplasty. In the absence of guiding evidence, rehabilitation visit usage varies considerably in the US, especially in the outpatient setting. We sought to develop expert consensus for visit usage recommendations in outpatient rehabilitation after total knee arthroplasty.

**Methods:** We used a 2-round Delphi process to develop expert consensus for visit usage recommendations. The Delphi panel consisted of surgeons, physical therapists, and advanced practice providers (N=29) with clinical and research expertise in recovery after total knee arthroplasty.

**Results:** The panel reached consensus on eight visit frequency recommendations for use in the first three months after total knee arthroplasty. These recommendations are responsive to patients’ time since surgery and their recovery status relative to expected. Twelve additional complementary recommendations were developed to be used with the visit frequency recommendations.

**Conclusions:** We used the Delphi process to develop expert recommendations for visit usage in outpatient rehabilitation after total knee arthroplasty. We envision these recommendations may help to facilitate a more preference-sensitive approach to postoperative rehabilitation.

## Introduction

No consensus recommendations exist for rehabilitation visit usage (i.e., visit timing, visit frequency, care duration) after total knee arthroplasty (TKA). Recent systematic reviews and clinical practice guidelines for post-TKA rehabilitation have provided no specific visit usage recommendations due to lack of supporting evidence—especially in the outpatient setting. ^1,2^ In the absence of guiding evidence, outpatient rehabilitation visit usage varies considerably in the US. Recent TKA rehabilitation studies have observed standard deviations ranging from 5-23 in the number of outpatient visits patients attend after TKA. ^3-7^ Warren et al. also found regional differences among Medicare beneficiaries in the average timing and duration of outpatient rehabilitation. ^8^ This practice variation may undermine the overall quality and cost-effectiveness of outpatient rehabilitation, ^9^ which is especially relevant given payors’ emphasis on reducing TKA costs. ^10,11^

Given the lack of available evidence, expert opinion could be useful for developing preliminary visit usage recommendations. In 2014, Westby et al. used the Delphi process to build expert consensus around best practices for TKA rehabilitation. ^12^ This study produced many key practice recommendations, but it did not develop consensus around visit usage in post-acute rehabilitation. The lack of consensus may have been partially due to (1) the wide scope of included rehabilitation topics, which might have limited the focus on visit usage and (2) the opinion among participants that rehabilitation should be individualized. ^12^

Individuals do have unique needs, goals, and expectations after TKA, ^13,14^ which suggests a preference-sensitive approach to visit usage may be ideal. ^15,16^ Preference-sensitive care occurs when well-informed patients make health care decisions in line with their individual preferences. ^16-18^ Some patients with TKA may prefer extensive rehabilitation to help them achieve ambitious goals, while others may prefer to recover mostly independently. However, because no evidence currently exists to help patients make informed decisions, visit usage is more likely driven by the capacity of local health care systems. ^17-20^ This type of supply-sensitive care fosters overutilization and ignores patients’ individual needs and preferences. Therefore, patients and clinicians could benefit from expert recommendations to anchor preference-sensitive decisions for visit usage in outpatient rehabilitation after TKA.

In this study, we used the Delphi method to develop expert consensus for visit usage recommendations after TKA. We focused specifically on the optimal visit timing, visit frequency, and duration of outpatient rehabilitation; we hypothesized this focused approach would avoid the challenges experienced in previous studies for developing consensus. Our overall goal was to create evidence that can be used to facilitate preference-sensitive visit usage decisions.

## Methods

### Panelist recruitment

We sought to enroll a heterogenous Delphi panel consisting of physical therapists, orthopedic surgeons, and advanced practice providers (e.g., physician assistants) with both clinical and research expertise in TKA recovery. We recruited participants from our own professional networks and through author lists of recently published literature in TKA rehabilitation. We limited our recruitment to individuals based in the United States because practice patterns can vary widely between countries. Individuals were eligible to participate if they had > 5 years of TKA-related experience and met one of the following volume criteria: (1) physical therapist who sees > 10 patients with TKA/year in the outpatient setting, (2) orthopedic surgeon who performs > 50 TKAs per year, or (3) advanced practice provider who sees > 50 patients with TKA/year. Additionally, clinicians from these professions who did not meet the volume criteria were eligible to participate if they had > 5 years of experience conducting and publishing TKA-related research. We aimed to enroll 30 participants with representation from each eligible profession; we chose this sample size as previous Delphi studies have observed stability with as few as 23 participants. ^21^ After enrollment, we sent participants personalized email reminders for each Delphi round to maximize response rates.

### Recommendation Development Phase

Before recruiting panelists, we developed a candidate list of visit usage recommendations using our collective expertise and existing TKA rehabilitation literature. We iteratively piloted these candidate recommendations and all survey materials with members of our research team and broader institutional network with relevant expertise in TKA rehabilitation. We used pilot feedback to revise recommendations, generate ideas for new recommendations, and ensure all survey materials were worded clearly and without bias. Using this process, we developed two categories of recommendations: (1) visit frequency recommendations and (2) complementary recommendations, which were designed to be used with the visit frequency recommendations. None of the individuals involved in the development phase participated on the Delphi panel.

### Visit frequency recommendations

We framed the visit frequency recommendations around efficient visit usage, which we defined as the minimum frequency of supervised visits needed to provide adequate care. We used two main strategies to develop these recommendations. First, we anchored each recommendation to a specific timeframe after surgery (i.e., postoperative month 1, 2, or 3). We used this strategy because visit frequency recommendations may depend on postoperative tissue healing. ^12^ Also, clinicians typically re-evaluate rehabilitation treatment plans on monthly intervals, and patients often discharge from rehabilitation within three months after surgery. ^3,7,8^ Second, we anchored each recommendation according to patients’ observed recovery status relative to their expected recovery (i.e., patient is demonstrating a slow, typical, or fast recovery relative to expected). We used this strategy because patients are expected to recover differently based on their individual characteristics, ^1^ and monitoring recovery against an expected value can be a useful decision making strategy. ^22^ Together, these strategies resulted in the development of nine separate combinations of recovery month + recovery status (e.g., postoperative month 1, recovering slower than expected). For each of these combinations, we asked panelists to rate their level of agreement with six different visit frequency options (0x/month, 1x/month, 2x/month, 1x/week, 2x/week, 3x/week). Overall, panelists considered 54 different visit frequency recommendations (nine combinations of recovery month + recovery status with six frequency options each). See Box 1 for an illustration of the structure used to create these recommendations.

#### Box 1. General structure used to create 54 different candidate visit frequency recommendations

|  |  |  |  |  |  |  |  |  |
| --- | --- | --- | --- | --- | --- | --- | --- | --- |
| Please rate your level of agreement or disagreement with the following visit frequency recommendations for outpatient rehabilitation frequency after TKA: |  |  |  |  |  |  |  |  |
| During the <u>first, second, or third</u><br>select one month after TKA, the most efficient visit frequency of outpatient rehabilitation for patients demonstrating a <u>typical, slow, or fast</u><br>select one recovery relative to expected should be <u>0x/month, 1x/month, 2x/month, 1x/week, 2x/week, or 3x/week</u><br>select one |  |  |  |  |  |  |  |  |
| **1**<br>**Strongly**<br>**Disagree** | **2** | **3**<br>**Somewhat**<br>**Disagree** | **4** | **5**<br>**Neutral** | **6** | **7**<br>**Somewhat**<br>**Agree** | **8** | **9**<br>**Strongly**<br>**Agree** |

### Complementary recommendations

We designed the complementary recommendations to be used with the visit frequency recommendations. They queried panelists’ opinions on (1) the optimal timing for initiating and stopping outpatient rehabilitation, (2) important range of motion recovery thresholds, (3) the approximate proportion of patients who demonstrate slow, typical, or fast recovery relative to expected, and (4) the safety and effectiveness of telerehabilitation for TKA.

#### Delphi Structure

We compiled the list of candidate recommendations into surveys for panelists to consider in Round 1 of the Delphi process. Panelists were asked to rate their level of agreement or disagreement with the visit frequency recommendations using a Likert scale ranging from 1 (strongly disagree) to 9 (strongly agree) (see Box 1). We used the RAND UCLA method to define consensus, ^23^ where recommendations with a median response of > 7/9 and less than 30% of responses in the 1-3 range were considered to have reached consensus. ^24,25^ Because panelists separately rated six different visit frequency options for each combination of recovery month + recovery status, more than one frequency within a combination could potentially meet our definition of consensus. In this case, we considered the frequency with the higher me an response to have reached consensus.

A few of the complementary recommendations were scored using the Likert scale and method described above, but most of them required numeric responses. We did not employ a formal definition of consensus for these numerically scored recommendations because they were meant to be supportive—not definitive. Instead, we calculated the mean response for these recommendations during the final Delphi round. In addition to Likert and numeric responses, the Delphi survey also included open-ended text boxes to record panelists’ comments after each recommendation. Panelists were encouraged to provide the rationale behind their response, their opinions on specific recommendations, and suggestions for new/revised recommendations.

We conducted additional rounds as needed to develop consensus. During each subsequent round, we included all previous recommendations that had not reached consensus, and we revised and added new recommendations based on panelist feedback. We also provided panelists with (1) a list of recommendations that previously reached consensus, (2) a comparison between the individual’s response and the group’s response for each recommendation, and (3) a qualitative summary of the group’s comments for recommendations from the previous round.

To determine whether additional rounds were needed, we examined the response stability for each recommendation that had not reached consensus. Specifically, we compared the absolute difference in the coefficient of variation (CV, standard deviation / mean) between rounds and considered values < 0.2 to be indicative of stability. ^26,27^ We also monitored the number and content of comments between rounds; we considered fewer comments with no major change in content as further evidence of response stability. ^28^ We discontinued the Delphi process once all recommendations had either reached consensus or demonstrated stability in the previous two rounds. See Figure 1 below for a summary of the Delphi round structure.

**Figure 1.**
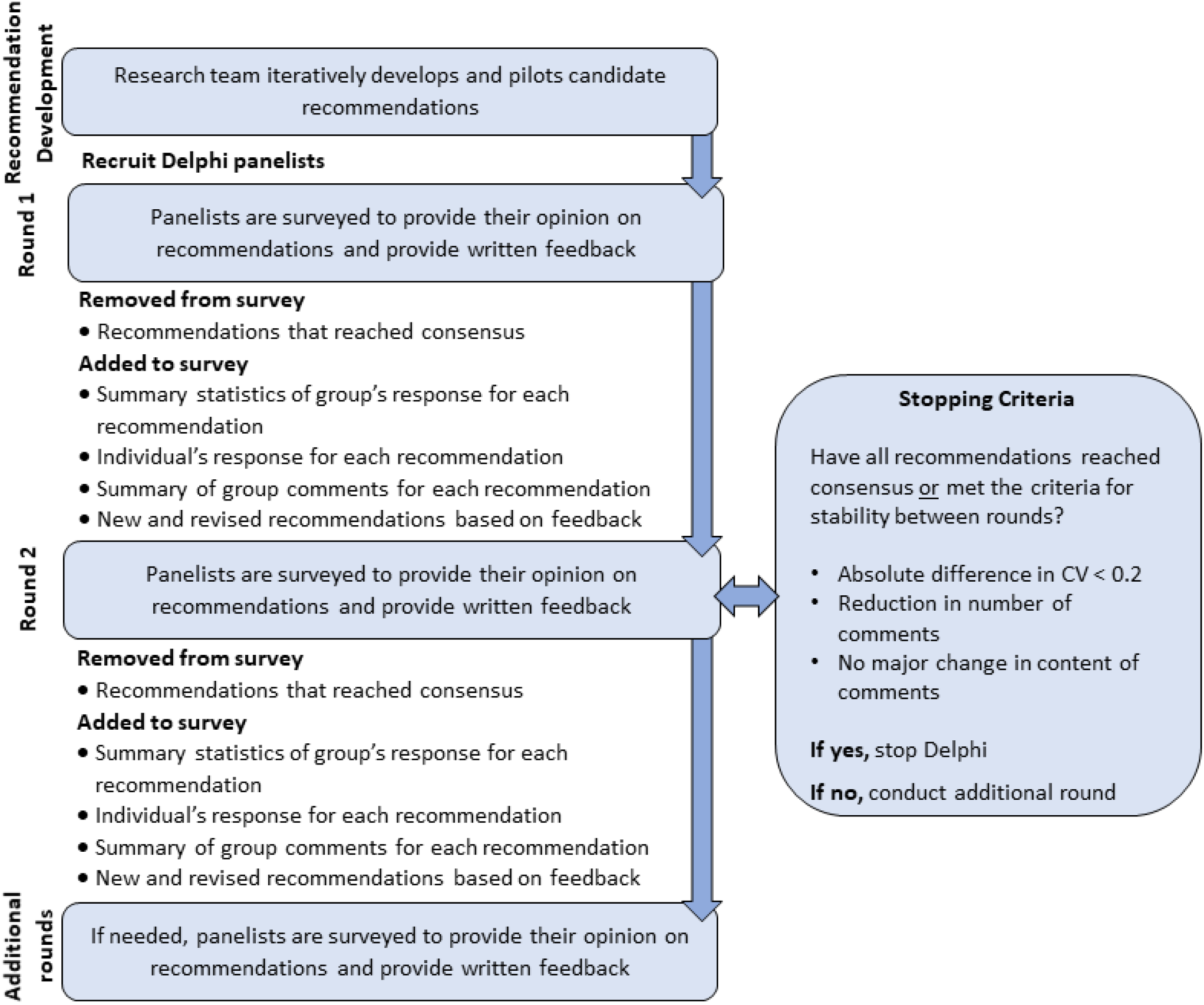
Visual summary of study structure

All study procedures were approved by the Colorado Multiple Institutional Review Board. We administered all surveys electronically via REDCap electronic data capture tools hosted at the University of Colorado. ^29,30^ We assured panelists their responses and identities would be anonymous throughout the study.

## Results

We invited 49 individuals to participate, and 30 panelists enrolled with 29 completing two Delphi rounds. The panel had a median of 17 years of TKA experience and included individuals from 11 US states and 22 unique zip codes. The panel included representation from the Department of Veterans Affairs (n=6), university settings (n=6), non-profit organizations (n=9), private practice (n=5), and other practice settings (n=3). Twelve out of the 18 participating physical therapists held either a PhD or board-certified specialization. Additional details regarding the panel’s experience and participation by round are available in Figure 2 and Table 1, respectively. We did not conduct additional rounds beyond Round 2 because all recommendations that had not reached consensus met our definition of stability between Round 1 and 2: (1) absolute difference in CV < 0.2, (2) reduced number of comments, (3) no major changes in content identified in comments.

**Figure 2.**
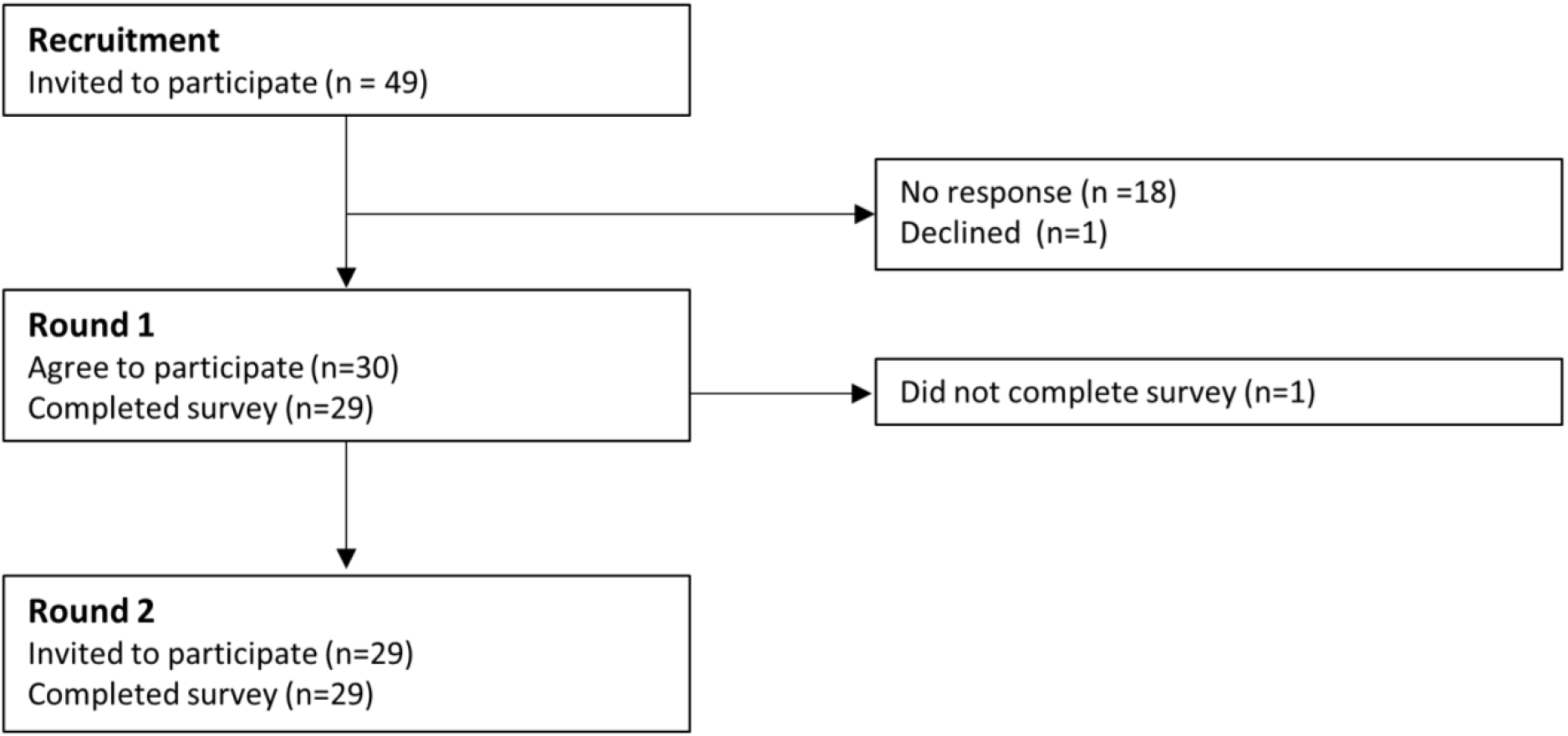
Flow chart depicting panelist recruitment and participation

**Table 1.** Summary of panel’s TKA-related experience. All values presented as median (range) unless otherwise noted

| Characteristic | Orthopedic surgeons (n=8) | Physical therapists (n=18) | Advanced practice providers (n=3) | Total (N=29) |
| --- | --- | --- | --- | --- |
| **Years of TKA experience** | 15.5 (11 – 36) | 15 (6 – 48) | 6 (6 – 18) | 15 (6 – 48) |
| **TKA experience** |  |  |  |  |
| Primarily clinical | n = 5 | n = 12 | n = 3 | n = 19 |
| Equally clinical + research | n = 3 | n = 4 | n = 0 | n = 8 |
| Primarily research | n = 0 | n = 2 | n = 0 | n = 2 |
| **Patients with TKA treated or operated on per year\*** | 250 (120 – 400) | 45 (12 – 180) | 240 (144 – 240) | 120 (12 – 400) |

\*Panelists with primarily research experience were not included in this calculation

**Table 2.**
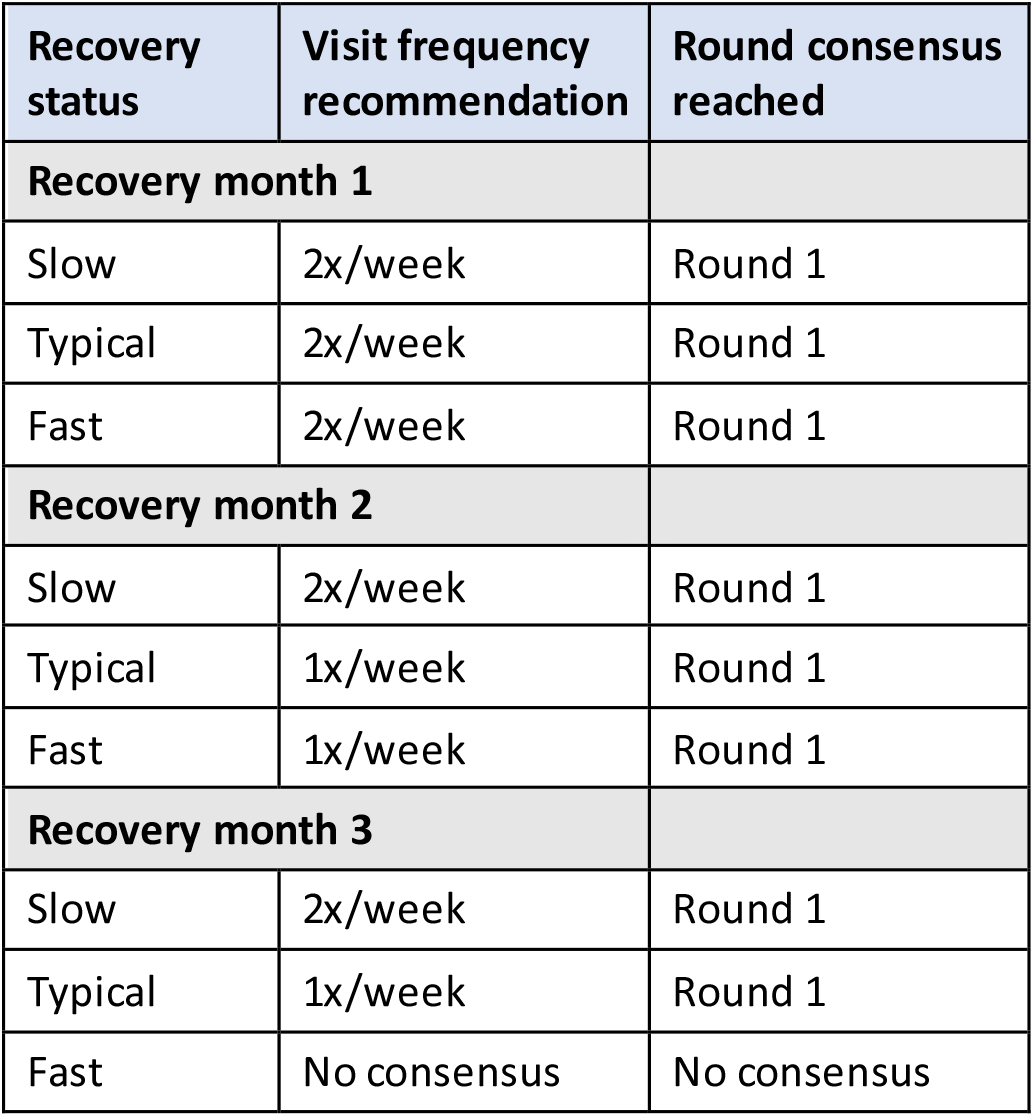
TKA rehabilitation visit frequency recommendations by patient’s postoperative month and recovery status

### Visit frequency recommendations for outpatient TKA rehabilitation

The panel reached consensus in Round 1 for visit frequency recommendations in eight of the nine combinations of recovery month + recovery status. The panel did not reach consensus on a recommendation for patients recovering faster than expected in postoperative month 3 during either round. In both rounds, many panelists commented that a fast-recovering individual’s need for rehabilitation at postoperative month 3 is highly contingent upon their postoperative goals. Other panelists commented that fast-recovering patients should already be discharged by postoperative month 3.

All complementary recommendations that used Likert scoring reached consensus. The panel agreed that outpatient rehabilitation should be initiated within 1 week following TKA, and that telerehabilitation is safe and effective for patients demonstrating a typical or fast recovery, but not for patients demonstrating a slow recovery. The panelists also provided stable numeric responses for (1) the optimal timing for discharge from outpatient rehabilitation, (2) the proportion of patients who demonstrate a slow, typical, or fast recovery, and (3) important knee flexion and extension range of motion thresholds. The complementary recommendations are displayed in Table 3 along with the format (Likert or numerical) used to score them.

**Table 3.** Complementary recommendations designed to be used with visit frequency recommendations

| Complementary recommendation | Scoring method (score) | Potential application |
| --- | --- | --- |
| **Timing of outpatient rehabilitation** |  |  |
| Assuming a patient does not receive sub-acute or home health rehabilitation, outpatient rehabilitation should be initiated within the first week after TKA | Likert<br>(Consensus reached in Round 1) | Clinicians may recommend patients to schedule their first visit within one week after surgery |
| On average, patients who are demonstrating a *slower recovery* than expected are anticipated to discharge from outpatient rehabilitation around 15 weeks after surgery | Numerical<br>(Mean = 15.2 weeks) | Clinicians may initiate discussions about discharge around these timeframes. Patients can use them to inform their preferences for when to discontinue supervised rehabilitation. |
| On average, patients who are demonstrating a *typical recovery* compared to expected are anticipated to discharge from outpatient rehabilitation around 10 weeks after surgery | Numerical<br>(Mean = 10.2 weeks) |  |
| On average, patients who are demonstrating a *faster recovery* than expected are anticipated to discharge from outpatient rehabilitation around 8 weeks after surgery | Numerical<br>(Mean = 7.5 weeks) |  |
| **Proportion of patients who demonstrate a slow, typical, or fast recovery** |  |  |
| Approximately 22% of patients demonstrate a *slower recovery* than expected after TKA, which may indicate they need more rehabilitation than anticipated | Numerical<br>(Mean = 22.4%) | Categorizing a patient's recovery requires clinical judgment. Clinicians can use these values to compare how frequently they are recommending more/less care than typical compared to expert opinion. |
| Approximately 54% of patients demonstrate a *typical recovery* compared to expected after TKA | Numerical<br>(Mean = 54.3%) |  |
| Approximately 24% of patients demonstrate a *faster recovery* than expected after TKA, which may indicate they need less rehabilitation than anticipated | Numerical<br>(Mean = 23.6%) |  |
| **Important range of motion thresholds** |  |  |
| Most patients need approximately 114 degrees of knee flexion range of motion to be functional with most daily activities after TKA | Numerical<br>(Mean = 113.6 degrees) | Certain tasks that most patients perform (e.g., walking) require a minimum range of motion. Patients who are not tracking |
| Most patients need to be within approximately 2 degrees of knee extension from neutral to be functional with most daily activities after TKA | Numerical<br>(Mean = 2.4 degrees) | towards these thresholds could be considered as slow recovering regardless of their recovery in other domains. |
| **Telerehabilitation** |  |  |
| Telerehabilitation *should not* be considered a safe and effective option for patients demonstrating a *slower recovery* than expected after TKA | Likert<br>(Consensus reached in Round 1) | Some patients may prefer telerehabilitation or have access to care challenges (transportation, rural area), which may influence their preference for rehabilitation usage. Clinicians may choose to offer telerehabilitation to patients if their recovery is progressing fast or typically. |
| Telerehabilitation *should* be considered a safe and effective option for patients demonstrating a *typical recovery* compared to expected after TKA | Likert<br>(Consensus reached in Round 1) |  |
| Telerehabilitation *should* be considered a safe and effective option for patients demonstrating a *faster recovery* than expected after TKA | Likert<br>(Consensus reached in Round 1) |  |

## Discussion

The panelists reached consensus on recommendations for the frequency, timing, and duration of outpatient rehabilitation during the first 3 months after TKA. We envision that patients and clinicians can use these expert recommendations as the starting point for preference-sensitive visit usage decisions. Their clinical utility may best be illustrated by example (see Box 2).

### Box 2. Example case of using recommendations to facilitate preference sensitive visit usage

Patient A was recently re-evaluated by their physical therapist 8 weeks after their TKA (start of month 3). Patient A is pleased with their recovery because their pain is much improved, and they have met their personal goal of returning to a walking program. Patient A’s physical therapist informs them their recovery has progressed typically thus far. Their physical therapist suggests that most patients are recommended to be seen 1x/week at this point after surgery, and discharge is typically recommended around week 10. Patient A considers this information with respect to their goals and preferences and decides to return for one additional visit at week 10. They would prefer to rehabilitate independently until then because they feel confident with their home exercise routine and have already achieved their primary goal.

The case above demonstrates a simplified example of how these recommendations could facilitate preference-sensitive care. Patient A used the best available evidence (expert opinion in this case) to make a decision about visit usage in line with their own preferences. ^19^ This type of preference-sensitive approach requires supporting evidence, which the results of this study provide, to ensure patients are well informed in their preferences. Conversely, supply sensitive care occurs in the absence of medical theory or evidence, and visit usage is driven primarily by the capacity of the local healthcare system. ^17-20^ Current post-TKA rehabilitation practice exemplifies supply-sensitive care, where visit usage varies considerably by clinician, facility, and region in the absence of guiding evidence. ^3-8^ Shifting towards a more preference-sensitive approach to rehabilitation could meaningfully improve patient outcomes; patients with knee osteoarthritis who make informed, preference-sensitive care decisions have reported higher quality of life, function, and satisfaction compared to patients who do not. ^31-33^ It may also reduce overall costs, ^17^ which could help ensure that outpatient rehabilitation remains a valued and reimbursable service after TKA.

The clinician’s role in facilitating preference-sensitive care using this study’s recommendations—or any medical evidence—should not be overlooked. To facilitate preference-sensitive care, clinicians must engage patients in the decision-making process, ^17^ attempt to maintain equipoise, ^16^ and avoid rigidly applying evidence. ^34^ Clinicians may face barriers to consistently practicing this way in outpatient rehabilitation after TKA. ^15^ Therefore, the recommendations from this study may be most effective when combined with clinician training^35^ or patient-facing support programs^36,37^ to facilitate preference-sensitive decisions for visit usage.

Clinicians must also use their clinical judgement when using the recommendations from this study. They need to determine whether a patient is demonstrating a slow, typical, or fast recovery relative to expected; the complementary recommendations were designed to aid in this determination. For example, a clinician may determine a patient’s recovery to be slower than expected if they are not on pace to achieve an important range of motion threshold (e.g., around 114 degrees of knee flexion based on this study’s results). Additional examples of how the complementary recommendations can be applied with the visit frequency recommendations are provided in Table 3. However, clinicians must consider numerous additional factors when assessing an individual’s recovery such as their preoperative prognosis, ^1^ pain, wound healing status, ^12^ and individual goals/expectations. ^13,14^ Clinicians and patients should work collaboratively to assess recovery and consider using these recommendations as a starting point for visit usage decisions. Clinicians should also ask patients about external factors that may influence their preferences such as familial support, access/transportation to rehabilitation, and insurance coverage. ^12,15^ Clinicians regularly make recommendations in consideration of these interplaying factors, but they should discuss their rationale with patients to facilitate preference-sensitive decisions.

The panelists in this study did not reach consensus on a visit frequency recommendation for individuals recovering faster than expected in postoperative month 3. It appeared unlikely that additional rounds would lead to consensus because panelists’ responses and comments were stable between rounds. The panelists appeared to be split among two groups based on their comments. One group felt these patients should already be discharged from rehabilitation, while the other group felt the recommended rehabilitation frequency for these patients should depend on the ambitiousness of their goals (which would be consistent with preference-sensitive care). Although no consensus was reached for a specific visit frequency, the complementary recommendations suggest that clinicians should consider discussing discharge with fast-recovering patients around 8 weeks after surgery.

This study does have a few limitations. Perhaps most notably, patients with TKA were not included on the Delphi panel. We chose not to include patients because we felt it was important for panelists to have experience with a wide range of cases given the heterogeneity of the TKA population. We also envisioned that an individual patient’s input on visit usage would be most valuable when applied to their own care (i.e., preference-sensitive care).

Regardless, future work should examine patients’ perceptions of the acceptability and usefulness of the recommendations developed in this study. This study also has notable strengths. We enrolled an experienced panel with considerable diversity in terms of profession, geography, and practice setting. To strengthen the validity of our findings, we used an iterative development phase, established predefined criteria for defining consensus and for stopping the Delphi process, provided participants with both quantitative and qualitative feedback between rounds, and incorporated panelist feedback into subsequent rounds. ^38,39^

## Conclusion

We used the Delphi method to develop recommendations foroutpatient rehabilitation usage after TKA. These recommendations can be used by clinicians and patients to facilitate a preference-sensitive approach to rehabilitation usage, which may improve the quality and efficiency of care.

## Data Availability

Data produced in the present study may be available upon reasonable request to the authors.

## Abbreviations

(TKA): total knee arthroplasty
(CV): coefficient of variation

